# Consensus on covert awareness: A Delphi study

**DOI:** 10.1101/2025.06.16.25329723

**Authors:** Caroline Schnakers, Berno Overbeek, Niko Fullmer, Liliana Teixeira, Matteo Zandalasini, Kseniia Yatsko, Ann-Marie Morrissey, Nathan Zasler, Anna Estraneo, IBIA Disorders of consciousness Special Interest Groups (DoC SIG)

**Author notes:** Corresponding author: Caroline Schnakers, PhD Associate Director, Research Institute, Casa Colina Hospital and Centers for Healthcare 255 East Bonita Avenue, Pomona, CA 91767 Phone: 909/596-7733, ext. 3038 •. Co-first authors (Caroline Schnakers, Berno Overbeek).

## Abstract

Background: Identifying willful brain activity in patients with Disorders of Consciousness (DoC) is critical, as some patients fail to exhibit behavioral signs of consciousness at the bedside but respond to active tasks via neuroimaging or electrophysiological measures. Standardized terminology for this subgroup is absent while it is essential for advancing research and clinical care. To determine the level of consensus among a large group of international experts on terminology and definitions for this clinical entity, as described by terms such as covert awareness (CA), cognitive motor dissociation (CMD), functional locked-in syndrome (fLIS), and non- behavioral minimally conscious state (MCS*). Methods: A Delphi study was conducted using REDCap to evaluate expert agreement on terminology and definitions. The study was conducted among international experts in DoC, primarily from Europe/UK, the USA, and other regions. ***Findings***: Ninety-six experts participated. Among these, 75 (78%) completed both rounds. Participants were predominantly clinical scientists (71%) working in rehabilitation settings (63%). A Delphi method was followed. Consensus on terminology and related definitions was defined as a median score of 5, an interquartile range (IQR) ≤ 1, and ≥ 75% agreement (scores of 4 or 5). Within two rounds, consensus was achieved for over two-thirds of the statements. The term "Covert Awareness" (CA) and its associated definition were identified as the preferred terminology by an international expert panel. ***Interpretation***: We recommend the use of "Covert Awareness" (CA) since our large group of international experts consistently agreed on such preferred term for this subgroup of patients with DoC. This consensus (>75% agreement) establishes a foundation both for future research and clinical standardization. The findings have implications for improving diagnostic accuracy and advancing understanding of covert awareness in DoC, although further study is needed to refine and apply the agreed-upon definition in clinical practice. ***Funding***: Nihil.

**Research in context:** 

**Evidence before this study:** Since the initial description of a patient with covert awareness in 2006 using mental imagery tasks during fMRI, a substantial number of publications has reported similar findings. Two meta-analyses have shown that 14–17% of patients diagnosed behaviorally as being in a vegetative state (VS) actually exhibit willful brain activity detectable through neuroimaging or electrophysiological paradigms. Recent multicenter studies reported that up to 25% of such patients may demonstrate covert command-following. Despite this growing body of evidence, the field has lacked a standardized nomenclature to describe this subgroup of patients. A 2022 systematic review identified 25 different terms in use—such as covert awareness (CA), cognitive motor dissociation (CMD), functional locked-in syndrome (fLIS), and non-behavioral minimally conscious state (MCS*)—underscoring the inconsistency in terminology. No prior study has evaluated expert consensus on preferred terminology and its definition.

**Added value of this study:** This study is the first international Delphi consensus effort specifically aimed at determining the most appropriate term and corresponding definition for patients with disorders of consciousness who exhibit willful brain activity detectable only through neuroimaging or electrophysiology. Thanks to a large group of international experts across diverse clinical and research backgrounds, consensus was reached on “Covert Awareness” (CA) as the preferred term, along with a precise definition. This consensus was robust across disciplines and geographic regions.

**Implications of all the available evidence:** Establishing consensus on the term “Covert Awareness” (CA) provides a critical foundation for future research, clinical practice, and guideline development. Standardized nomenclature can improve diagnostic consistency, enhance communication across care teams, and promote better public and caregiver understanding. It also enables more rigorous comparative research by providing clear inclusion criteria for future studies. While additional refinement of the definition may be required, this consensus marks a key step in formalizing the recognition of CA as a distinct clinical entity. Future efforts should also consider the development of specialized diagnostic centers and the integration of CA detection into clinical pathways and treatment trials.

## Introduction

Detecting behavioral signs of consciousness in patients with severe acquired brain injuries can be challenging as it can lead to a misdiagnosis in approximately 40% of cases when no standardized assessment tool is used.^1^ Even with the most careful behavioral assessment, willful brain activity might still be missed in some patients with disorders of consciousness (DoC) due to severe motor disabilities.^2^ For the past two decades, clinicians have been facing a subgroup of patients who are unable to show any behavioral sign of consciousness at the bedside but are able to respond mentally to neuroimaging (i.e., functional Magnetic Resonance Imaging – fMRI) or electrophysiological active task paradigms. The first report of this phenomena was published in 2006 and described the case of a young woman with severe brain injury diagnosed as being in a Vegetative State or Unresponsive Wakefulness Syndrome (VS/UWS; i.e., awake but no evidence of willful behaviors).^3–4^ When performing a mental imagery task (e.g., imaging playing tennis), her fMRI-related brain activity was similar to those observed in healthy controls.^5^ Later, in a study using the same fMRI paradigm in a bigger sample (n=54), two patients clinically diagnosed as being in a VS/UWS and three patients clinically diagnosed as being in a Minimally Conscious Sate (MCS; inconsistent but reproducible conscious behaviors)^6^ were able to perform the task. Moreover, one of these patients was able to correctly answer ‘yes’ or ‘no’ to autobiographical questions by using either motor or spatial imagery.^7^ Since then, this condition has been extensively reported in the literature. Two meta-analyses found that consciousness can be observed in 14-17% of patients whose behavioral assessment suggested VS/UWS.^8–9^ A recent multicentric study revealed a rate as high as 25% of these patients being able to follow instructions.^10^ The existence of this unique population led the American Academy of Neurology and the European Academy of Neurology to strongly recommend using complementary techniques such as neuroimaging and electrophysiology when diagnosing patients with DoC.^11–12^

Even though researchers and clinicians recognize the existence of this clinical entity, there is still no consensus regarding the nomenclature to describe it.^13^ A recent systematic review found 25 existing names for this condition, with several of these names sometimes used interchangeably within the same manuscript. The names most frequently used included covert awareness (CA), cognitive motor dissociation (CMD), functional locked-in syndrome (fLIS), and non-behavioral MCS (MCS*).^14^ Each of these terms have been defined in different ways in the existing literature calling for a consensus on the best clinical label for the condition. Indeed, using a clear taxonomy, along with precise definitions, has several key benefits. It supports rigorous research among scientists, facilitates effective communication and diagnostic consistency among healthcare professionals, and upholds trust and professionalism when interacting with families and the broader public. ^15^ A precise taxonomy would therefore be helpful for both scientists and clinicians.

In this context, the present Delphi study aimed to examine the level of agreement among a large group of international experts on these terms and their related definitions across the available literature. The overarching goal of our study was to inform future discussions on taxonomy for this condition.

## Methods

Based on an extensive literature review,^14^ a Delphi method was used to investigate the level of consensus on names and related definitions for CA, CMD, fLIS, and MCS*. A Delphi study is a multi-staged survey, which aims to achieve consensus on an important issue.^16^ The result is an expert opinion about a subject where previously no such opinion existed. In the development of clinical guidelines, the Delphi method is used as a consensus building process to capture expert opinions and experiences.^11,17^

### Survey development

Since international consensus was aimed for, a core group of five members from the Special Interest Group on DoC of the International Brain Injury Association (IBIA DoC SIG) was formed. The members of this group are all experts in their respective country (USA, the Netherlands, Ireland, Russia, Portugal and Italy) and participated in previous projects on the same topic.^9,14^ Additionally, one member (BO) had previous experiences with using the Delphi method.^18^ The core group developed the survey but did not participate in the study. Survey data were collected and managed using Research Electronic Data Capture (REDCap) hosted at Casa Colina Hospital and Centers for Healthcare. REDCap is a secure, web-based software platform designed to support data capture for research studies.^19^

The survey included, across rounds, 11 to 16 statements related to definitions and two to three questions related to name preferences. For each statement, the level of agreement was indicated on a five-point Likert scale as used in Delphi studies:^16^ strongly disagree (1), moderately disagree (2), neutral (3), moderately agree (4), and strongly agree (5). A comment section was also included for each definition and was used for additions and/or revisions of statements in the second round.

The statements used for each definition were based on a previous systematic review^14^ that was accessible to each participant using a direct link to the article embedded in the survey. Before starting the Delphi process, a pilot was done among core members to test the survey and estimate completion time. The survey duration was kept less than 30 minutes to decrease risks of incomplete surveys or drop-outs (eTable1).

### Expert criteria

After the core group approved the final version of the survey, an international panel of experts were identified and selected using one or more/one of the following criteria: (1) at least 5 years of research and/or clinical experience in the assessment and treatment of patients with a DoC, in the acute, post-acute or chronic phases, (2) published at least 5 peer-reviewed articles on DoCs over the last 5 years. The core group agreed that a large sample would be needed to optimize the reliability (less influence of outliers, more resilience to dropouts) as well as the generalizability of our results. Three hundred experts were contacted, based on rate of participation (45%) and loss-to-follow up (25%) in a previous Delphi study involving experts in DoC.^18^ To reach this high number of potential experts, participants were recruited by contacting IBIA DoC SIG members and by snowball sampling.

### Data Collection

Based on the Delphi method, ^16,20^ the core group agreed on a two to three round of data collection, since this approach provides the best balance between obtaining a consensus and maintaining a high response rate. The core group also agreed that a third round would not be performed if consensus was reached for more than two thirds of the statements on Round 2 and that a maximum of three rounds would be done even if consensus for agreement could not be reached for more than two thirds of these statements.^16,20^ Each identified expert received an e-mail invitation including an individualized link, gave digital informed consent before starting the first-round and were then given access to the survey. Responses were anonymized in REDCap; all responders being related to a number upon starting the survey. Demographic data were collected in Round 1 to document the participants’ profile and to ensure a fit with our criteria for expertise (i.e., country of residence, profession, experience working with DoCs in years, number of relevant publications, work setting, use of assessment scales and use of additional diagnostic techniques). Participants had the possibility to “save and return later” after starting the survey to prevent fatigue among responders (i.e., potential consensus bias). After submitting feedback, each participant received an email with a pdf summarizing their responses, which they were asked to review just before answering next round of the survey. Each round was open for 3 weeks (with an automatic reminder after 7 days) and had a 1-week (Round 2) to 2 weeks (Round 1) interval in-between rounds to allow time for analysis. The analyses were done by the core group. The Delphi study took place between the beginning of April until the end of May 2024.

### Statistical analysis

Based on the Delphi method,^16,20^ measures of central tendency and dispersion were used. Consensus was also calculated by combining median values, interquartile ranges (IQR), and percentage of agreement/disagreement. As done in most Delphi studies, consensus for agreement was defined as a median score of 5, an IQR ≤ 1, and ≥ 75% scoring 4 or 5 while consensus for disagreement was defined as a median of 1, an IQR ≤ 1, and ≥ 75% scoring 1 or 2. ^16,20^ If consensus for agreement (or disagreement) was reached for a statement, this statement was not presented again in the next round.

The protocol was reviewed and approved by the Institutional Review Board (IRB) committee of Casa Colina Hospital and Centers for Healthcare, Pomona, CA, USA.

## Results

### Expert panel

Ninety-six experts agreed to participate and fit our criteria for expertise. Seventy-five experts (78%) completed all rounds (Fig. 1). Most experts were clinical scientists (i.e., medical experts who study new techniques to help prevent, diagnose and treat illness) (71%). Participants worked as medical doctors (51%; specialized in neurology, intensive care medicine, neurosurgery, and, physical medicine and rehabilitation), psychologists (13%), physical therapists (5%), occupational therapists (5%), speech therapists (3%), registered nurses (2%) as well as (neuro)scientists (20%). Most experts (64%) worked in a rehabilitation setting, 45% in an acute setting, 23% in a long-term care setting, and 8% in a research setting. Forty nine percent were located in Europe and United Kingdom (EU/UK), 32% in the United States of America (USA) and 19% from other parts of the world (Others; i.e., Russia, Canada, Australia, Israel, Brazil, Malaysia and China). When diagnosing DoCs, behavioral assessments were the most frequently used (90%) by these experts, followed by electrophysiology (67% electroencephalogram-EEG and 42% event-related potentials-ERP), neuroimaging (34% fMRI, 27% positron emission tomography-PET, 7% single-photon emission computed tomography-SPECT). Other techniques such as TMS-EEG (21% - combining transcranial magnetic stimulation and EEG) or Brain Computer Interface (BCI; 13%) were also used. Active paradigms were less frequently used (28%) than resting-state (48%) or passive paradigms (37%) (Table 1).

**Figure 1.**
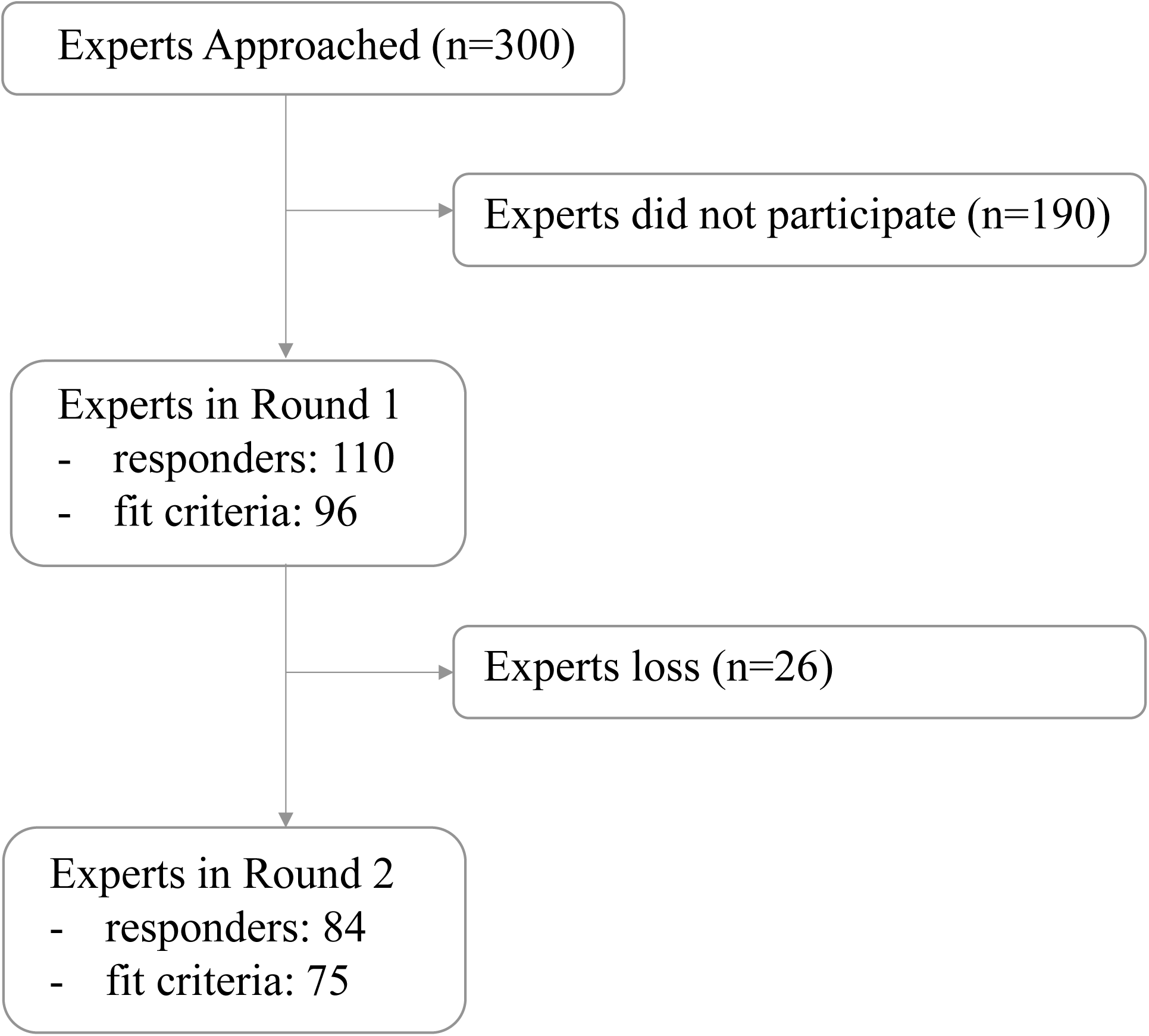
Flow chart of participation of experts

**Table 1.**
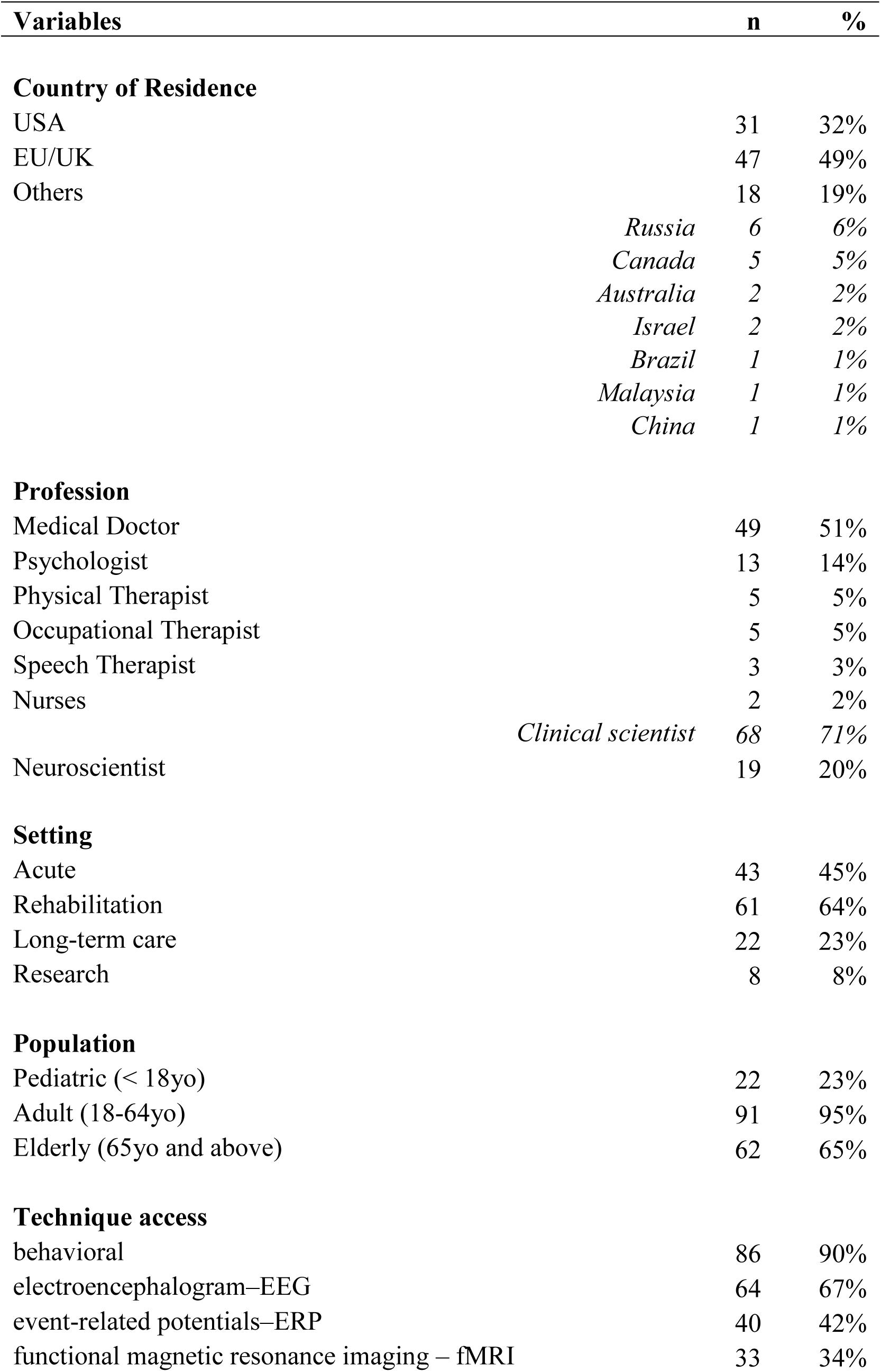
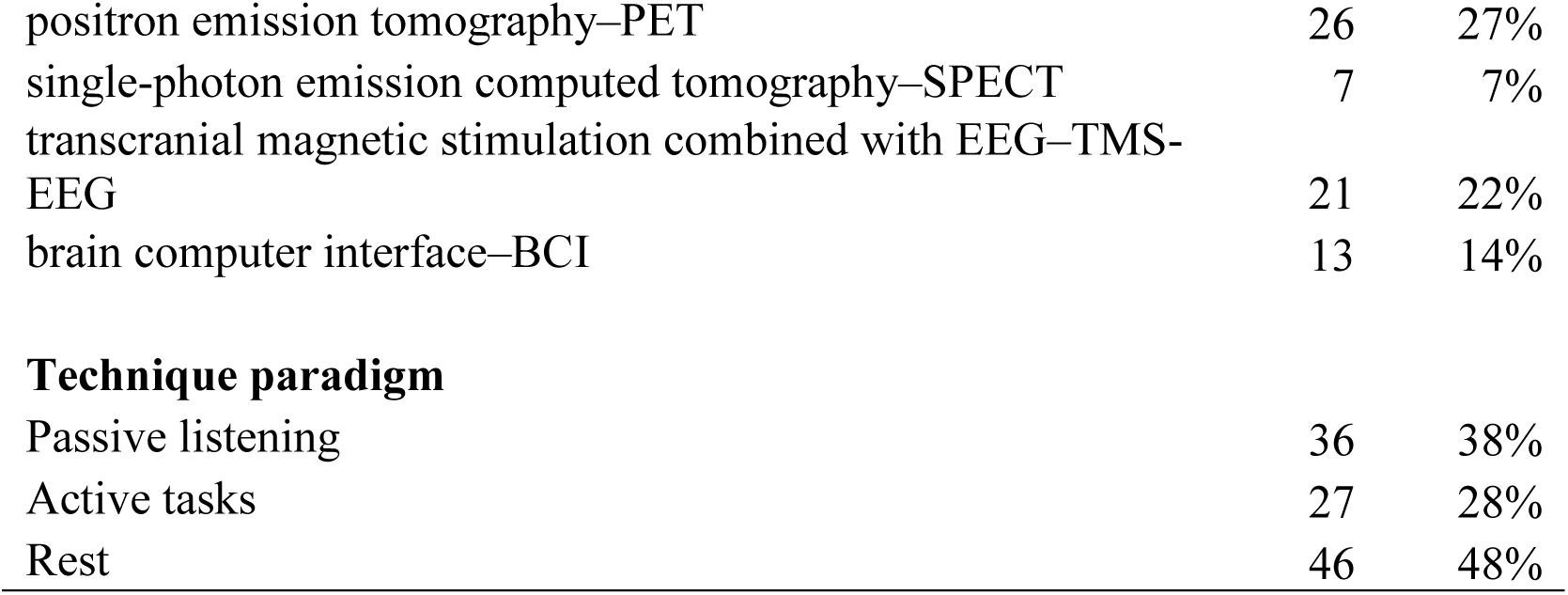
Participants characteristics.

### Consensus development

This Delphi study included two rounds. Indeed, a third round was not necessary since consensus was reached for more than two thirds of the statements on Round 2 (cf. Delphi methodology described above). ^16,20^

*Round 1.* Consensus was only reached for 5 out of the 16 (31%) definition statements (2/5 for CA, 1/3 for CMD, 2/5 for fLIS, and 0/3 for MCS*) (Table 2). Also, no consensus was reached on questions related to name preferences (Table 3). For the definition statements, all statements that did not reach consensus were rephrased for Round 2 based on the experts’ feedback. For name preferences, one question related to the choice of a covert related name (i.e., covert awareness, covert consciousness, covert cognition) was also included in Round 2 based on this feedback.

**Table 2.**
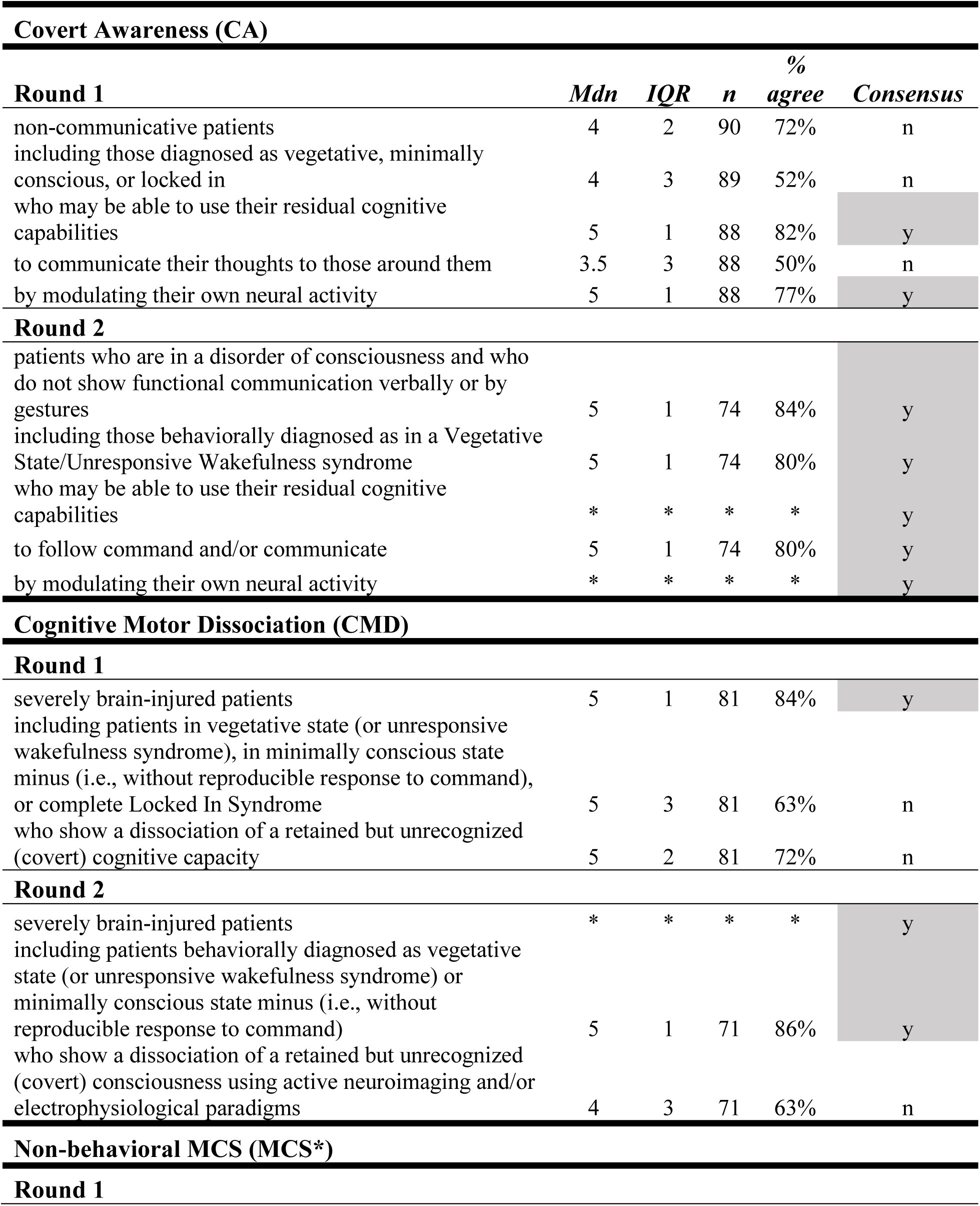
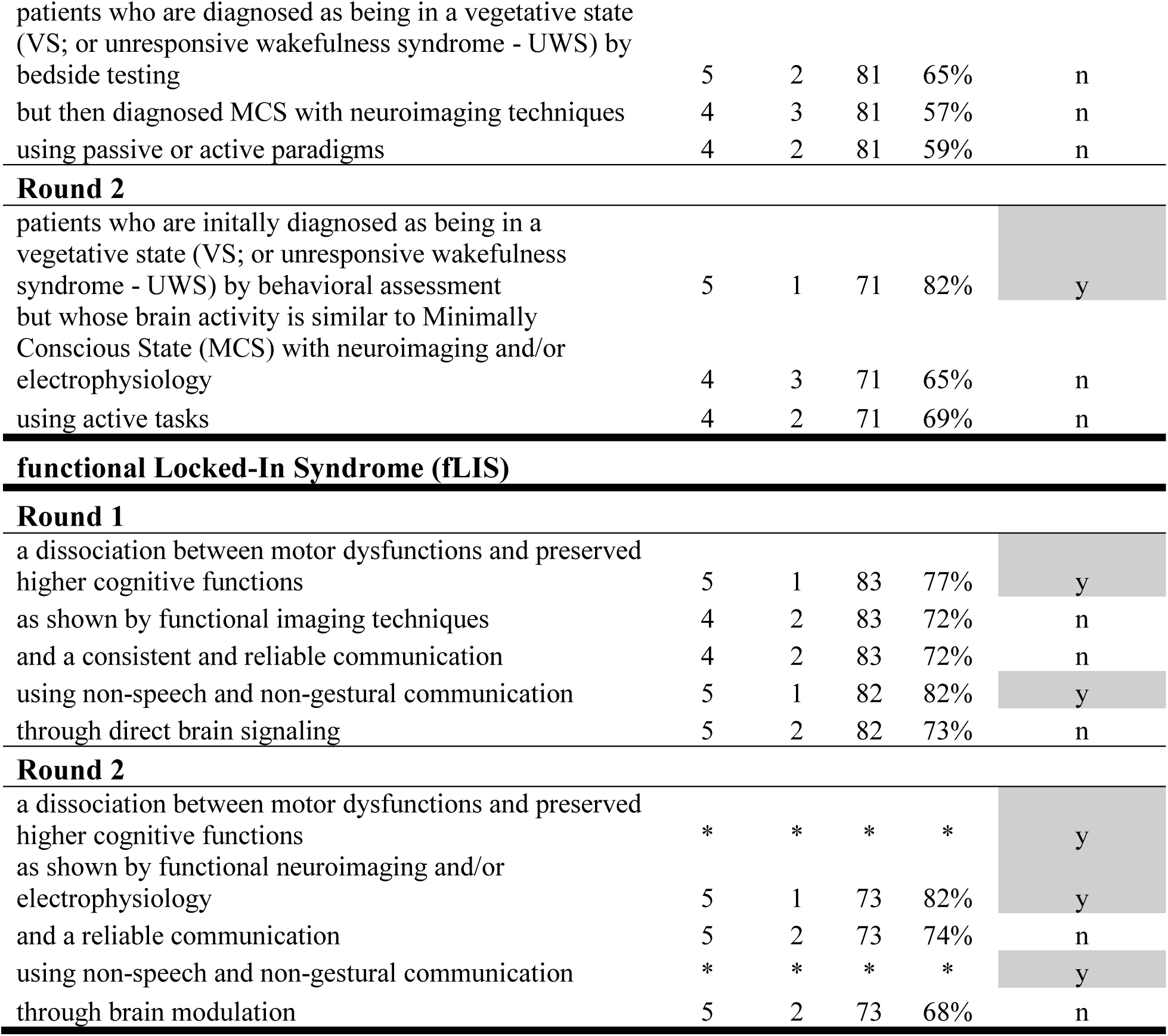
Statements with agreement rates in the round 1 and 2.

| **Covert Awareness (CA)** |  |  |  |  |  |
| --- | --- | --- | --- | --- | --- |
| **Round 1** | *Mdn* | *IQR* | *n* | *% agree* | *Consensus* |
| non-communicative patients | 4 | 2 | 90 | 72% | n |
| including those diagnosed as vegetative, minimally conscious, or locked in | 4 | 3 | 89 | 52% | n |
| who may be able to use their residual cognitive capabilities | 5 | 1 | 88 | 82% | y |
| to communicate their thoughts to those around them | 3.5 | 3 | 88 | 50% | n |
| by modulating their own neural activity | 5 | 1 | 88 | 77% | y |
| **Round 2** |  |  |  |  |  |
| patients who are in a disorder of consciousness and who do not show functional communication verbally or by gestures | 5 | 1 | 74 | 84% | y |
| including those behaviorally diagnosed as in a Vegetative State/Unresponsive Wakefulness syndrome | 5 | 1 | 74 | 80% | y |
| who may be able to use their residual cognitive capabilities | * | * | * | * | y |
| to follow command and/or communicate | 5 | 1 | 74 | 80% | y |
| by modulating their own neural activity | * | * | * | * | y |
| **Cognitive Motor Dissociation (CMD)** |  |  |  |  |  |
| **Round 1** |  |  |  |  |  |
| severely brain-injured patients | 5 | 1 | 81 | 84% | y |
| including patients in vegetative state (or unresponsive wakefulness syndrome), in minimally conscious state minus (i.e., without reproducible response to command), or complete Locked In Syndrome | 5 | 3 | 81 | 63% | n |
| who show a dissociation of a retained but unrecognized (covert) cognitive capacity | 5 | 2 | 81 | 72% | n |
| **Round 2** |  |  |  |  |  |
| severely brain-injured patients | * | * | * | * | y |
| including patients behaviorally diagnosed as vegetative state (or unresponsive wakefulness syndrome) or minimally conscious state minus (i.e., without reproducible response to command) | 5 | 1 | 71 | 86% | y |
| who show a dissociation of a retained but unrecognized (covert) consciousness using active neuroimaging and/or electrophysiological paradigms | 4 | 3 | 71 | 63% | n |
| **Non-behavioral MCS (MCS\*)** |  |  |  |  |  |
| **Round 1** |  |  |  |  |  |
| patients who are diagnosed as being in a vegetative state (VS; or unresponsive wakefulness syndrome - UWS) by bedside testing | 5 | 2 | 81 | 65% | n |
| but then diagnosed MCS with neuroimaging techniques | 4 | 3 | 81 | 57% | n |
| using passive or active paradigms | 4 | 2 | 81 | 59% | n |
| Round 2 |  |  |  |  |  |
| patients who are initially diagnosed as being in a vegetative state (VS; or unresponsive wakefulness syndrome - UWS) by behavioral assessment | 5 | 1 | 71 | 82% | y |
| but whose brain activity is similar to Minimally Conscious State (MCS) with neuroimaging and/or electrophysiology | 4 | 3 | 71 | 65% | n |
| using active tasks | 4 | 2 | 71 | 69% | n |
| Round 1 |  |  |  |  |  |
| a dissociation between motor dysfunctions and preserved higher cognitive functions | 5 | 1 | 83 | 77% | y |
| as shown by functional imaging techniques | 4 | 2 | 83 | 72% | n |
| and a consistent and reliable communication | 4 | 2 | 83 | 72% | n |
| using non-speech and non-gestural communication | 5 | 1 | 82 | 82% | y |
| through direct brain signaling | 5 | 2 | 82 | 73% | n |
| Round 2 |  |  |  |  |  |
| a dissociation between motor dysfunctions and preserved higher cognitive functions | * | * | * | * | y |
| as shown by functional neuroimaging and/or electrophysiology | 5 | 1 | 73 | 82% | y |
| and a reliable communication | 5 | 2 | 73 | 74% | n |
| using non-speech and non-gestural communication | * | * | * | * | y |
| through brain modulation | 5 | 2 | 73 | 68% | n |

**Table 3.** Results related to the question: “To what extent do you agree with the following name(s)?”

| **Round 1** | ***Mdn*** | ***IQR*** | ***n*** | ***% agree*** | ***Consensus*** |
| --- | --- | --- | --- | --- | --- |
| **Before taking the survey** |  |  |  |  |  |
| Covert Awareness (CA) | 4 | 1 | 95 | 78% | n |
| Cognitive Motor Dissociation (CMD) | 4 | 2 | 95 | 69% | n |
| Non-behavioral MCS (MCS*) | 3 | 2 | 95 | 35% | n |
| functional Locked-In Syndrome (fLIS) | 2 | 2 | 95 | 22% | n |
| **After taking the survey** |  |  |  |  |  |
| Covert Awareness (CA) | 4 | 1 | 81 | 77% | n |
| Cognitive Motor Dissociation (CMD) | 4 | 2 | 81 | 67% | n |
| Non-behavioral MCS (MCS*) | 3 | 2 | 81 | 44% | n |
| functional Locked-In Syndrome (fLIS) | 2 | 3 | 81 | 26% | n |
| **Round 2** |  |  |  |  |  |
| **Before taking the survey** |  |  |  |  |  |
| Covert Awareness (CA) | 5 | 1 | 75 | 95% | y |
| Cognitive Motor Dissociation (CMD) | 4 | 2 | 75 | 65% | n |
| Non-behavioral MCS (MCS*) | 2 | 3 | 75 | 27% | n |
| functional Locked-In Syndrome (fLIS) | 1 | 1 | 75 | 8% | y |
| **After taking the survey** |  |  |  |  |  |
| Covert Awareness (CA) | 5 | 1 | 71 | 87% | y |
| Cognitive Motor Dissociation (CMD) | 4 | 2 | 71 | 73% | n |
| Non-behavioral MCS (MCS*) | 2 | 3 | 71 | 30% | n |
| functional Locked-In Syndrome (fLIS) | 2 | 2 | 69 | 12% | n |
| **Covert related names** |  |  |  |  |  |
| Covert awareness | 5 | 1 | 71 | 83% | y |
| Covert consciousness | 4 | 2 | 71 | 66% | n |
| Covert cognition | 3 | 2 | 71 | 34% | n |

*Round 2.* Consensus for agreement was reached for 11 out of the 16 (69%) statements (5/5 for CA, 2/3 for CMD, 3/5 for fLIS, and 1/3 for MCS*). A full consensus was reached on a definition for CA (Table 2). Moreover, a consensus only for CA was reached when experts were asked which name they would agree upon (87%), and which covert-related names they would prefer (83%). A consensus for disagreement was reached on fLIS in Round 2 for questions related to name preferences (84%) (Table 3). When considering background and only selecting neuroscientist experts, same results were found; i.e., a consensus for all statements of the definition for CA only and a consensus on CA as the name preferred for this condition (over CMD, MCS* and fLIS as well as over covert consciousness or covert cognition) (Fig. 2; eTable 2 and 3). The consensus was also compared among geographic locations (USA vs. EU/UK vs. Others, eTable 2 and 3). A consensus on CA as the name preferred for this condition was consistently found (over CMD, MCS* and fLIS) across all locations. Compared to other covert-related names (such as covert consciousness or covert cognition), the highest agreement was observed for the term CA (>75%) but a consensus was not reached among USA and EU/UK responders. Regarding CA definition, a consensus was reached for all statements among responders of EU/UK vs. Others, while two statements did not reach consensus among USA responders (Fig. 2; eTable 2 and 3).

**Figure 2.**
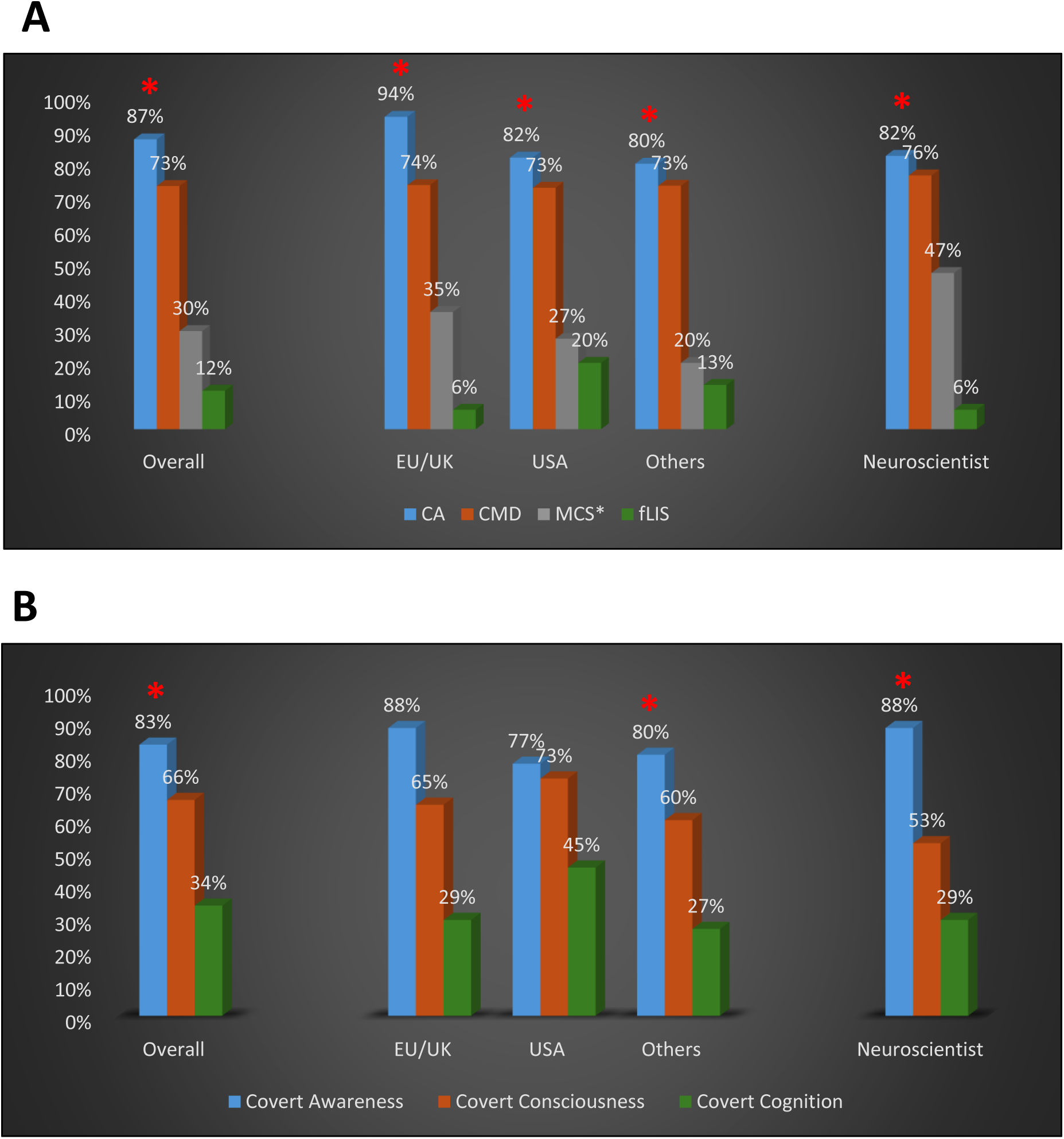
Consensus (*) overall, according to geographic location (EU/UK, USA, Others) and background (neuroscientist) for CA, CMD, MCS*, fLIS (A) as well as for covert related names (B) ***Legend***: covert awareness (CA), cognitive motor dissociation (CMD), functional locked-in (fLIS), and non-behavioral MCS (MCS*)

## Discussion

This international study examined the level of agreement on the nomenclature and related definitions for a clinical entity that was identified almost 20 years ago^5^ and has received various names through the years without any consensus among experts.^14^

Even though this study did not aim to reach a consensus on such a challenging topic, our results show that the term CA (i.e., “covert awareness”) quickly reached a consistent consensus (>75% agreement) among the large group of international experts involved in this study (96 in Round 1 and 75 in Round 2). A consensus was also reached on its definition; i.e.: “patients who are in a disorder of consciousness and who do not show functional communication verbally or by gestures, including those behaviorally diagnosed as in a VS/UWS, but who may be able to use their residual cognitive capabilities to follow command and/or communicate by modulating their own neural activity”.

Most of our experts were clinical scientists (71%). Since this background was over-represented, secondary analyses were performed focusing on a subgroup of experts with a neuroscience background. These analyses showed consistent results with a consensus on CA definition and on CA as the name preferred for this condition (over CMD, MCS*, fLIS, covert consciousness or covert cognition). Geographic locations were also compared and showed slightly different results. A consensus across locations was reached on CA as the preferred name over CMD, MCS* and fLIS but, despite CA receiving the highest agreement (>75%), a consensus was not reached on CA as the preferred name over covert consciousness or covert cognition among USA and EU/UK responders. Finally, a consensus for all statements of CA definition was observed among responders of EU/UK and Others but not among USA responders with two of the statements not reaching consensus.

Despite slight differences, our results show a consistent result across backgrounds and locations: a consensus on CA as the preferred name over CMD, MCS* and fLIS. The term CA was originally used by Owen and his collaborators (located in the UK) in one of the first papers on the topic^21^ and is one of the most highly referenced in the literature.^14^ However, such a quick consensus was rather surprising since “Cognitive Motor Dissociation” or CMD is frequently used in the DoC literature. This name was first mentioned by Schiff and coworkers (located in USA) in 2015 in response to a publication by Fernández-Espejo and coworkers (2015), who showed structural disconnection between thalamus and primary motor cortex in one “covertly aware” patient.^22^

Schiff and coworkers wrote an editorial on these results and suggested the term CMD to account for “…the sharp dissociation of a retained but unrecognized (covert) cognitive capacity in some severely brain-injured patients with non-purposeful or absent behavioral responses”.^23^ This term has been used in recent high-profile publications^10,24^ suggesting it was adopted by the field. To the contrary, the current Delphi study demonstrates otherwise as no consensus was reached on the name CMD and its definition. Based on the experts’ feedback, most of the debate was about: 1) the vagueness of the definition, which could apply to completely unrelated pathologies such as LIS^25^ or Amyotrophic Lateral Sclerosis (ALS)^26^ and; 2) more importantly, the fact that such a name implies that cognition is intact (which is currently unknown) and which could lead to misunderstanding among researchers, healthcare professionals or when interacting with families and the broader public. Other terms such as MCS* and fLIS failed to reach consensus. Most experts criticized MCS* for the inclusion of a passive paradigm to detect covert awareness which do not assess willful brain activity and might lead to false positives. The term “functional locked-in” or fLIS was the only term that reached consensus for disagreement as most experts found it confusing and disliked the direct comparison with (classical/incomplete) LIS that has well defined neuropathology and a different clinical profile.^25^

Our results also showed that, when diagnosing DoCs, techniques such as EEG (67%) are more frequently used by clinical experts as compared to functional neuroimaging (fMRI, 34%). Active paradigms that are usually involved in detecting willful brain activity are used in a minority of cases (28%) as compared to resting state (48%), which is consistent with prior findings.^27,28^ This reflects the reality of clinical versus research settings. Indeed, even though it is possible to use cheaper bedside techniques such as EEG in most clinical settings, it is challenging to use active paradigms as it requires a level of expertise often unavailable in non-academic settings. These challenges have been previously highlighted in the literature and call for the development of centers of expertise that clinicians could refer to, when implementing paradigms and analyzing their data.^24,29^ This is particularly needed since a recent multicentric study including 353 patients found that 25% of patients without an observable response to commands (and 20% of patients diagnosed in coma and VS/UWS) demonstrated covert awareness.^10^ Moreover, not only diagnostic techniques but also treatment strategies should be developed and/or implemented (e.g., using BCI or neuromodulation techniques) for a condition identified almost 20 years ago. However, for research to go further, we not only need a better understanding of the biomarkers related to this condition^24,30^ but also, and foremost, a clear taxonomy to allow a better communication between experts.

This study nevertheless has some limitations. Our results were stable across background, when comparing clinical scientists to neuroscientists. However, it would be interesting to confirm our findings in a bigger sample of neuroscientists to ensure reliability since the sample included in this study was relatively small (n=17). Also, part of our responders was from non-native English-speaking countries which could have affected our findings. Our results were comparable for the preferred name (CA) across locations (e.g., USA, EU/UK, Others) but the level of agreement for definition statements related to CA differed across locations. Further work might therefore be needed on the definition of CA itself in the future. Third, one could criticize the fact that definitions related to other covert-related terms were not included in this study. Indeed, based on an extensive literature review,^14^ CA was chosen since it was the most frequent term used in the literature, among all covert-related terms. Moreover, when other covert-related names (i.e., covert-consciousness and covert cognition) were included in Round 2, a consensus only on CA was still obtained. In any case, future discussions should address in which context CA should be applied and whether the use of other terms should be avoided or applied to distinct, separate situations. For example, one could imagine that while CA would apply to a VS/UWS who respond to command using active neuroimaging tasks, CMD might apply to patients who reliably communicate using such tasks and who should therefore be considered out of DoC (cf. Aspen criteria for the emergence from MCS).^6^ Finally, an interesting aspect that has not been tackled in this study is the opinion of caregivers of patients with DoC. Ensuring that a chosen taxonomy is accepted by experts and non-experts is the ultimate step since such a choice can impact not only caregivers but also public understanding of this condition.

In conclusion, based on our results, we recommend the use of "Covert Awareness" (CA) since our large group of international experts consistently agreed on such term for this subgroup of patients with DoC. This consensus, based on high agreement (>75%), establishes a foundation both for future research and clinical standardization.^31^ The findings have implications for improving diagnostic and prognostic accuracy as well as advancing understanding of covert awareness in DoC, although further study is needed to refine and apply the agreed-upon definition in clinical practice.

## Supporting information

eTable123

## Data Availability

Deidentified participant data will be accessible upon request to researchers whose proposed data use has received approval from an ethical committee for a specified purpose. Access will be granted by the corresponding author following the completion of a signed data access agreement.

## Acknowledgments

CS, BO, NF, LT, MZ, KY, AMM made substantial contributions to conception or design of the work, or the acquisition, analysis, or interpretation of data for the work; CS, BO, LT, MZ, KY, AMM, NZ, AE have drafted the work or reviewing it critically for important intellectual content; All authors have given final approval of the version to be published. CS agrees to be accountable for all aspects of the work in ensuring that questions related to the accuracy or integrity of any part of the work are appropriately investigated and resolved. None of the authors have financial or personal relationships or affiliations that could influence or bias their decisions, work, or manuscript. We have no source of funding to declare for this work.

