## Supplementary material for "Consensus on covert awareness: A Delphi study": eTable123

**Online-Only Supplements**

**eTable 1.** Survey questions for Rounds 1 and 2

| **Round 1** | |
|---|---|
| **Country of Residence** | |
| **Profession** | Medical Doctor |
| | Psychologist |
| | Physical Therapist |
| | Occupational Therapist |
| | Speech Therapist |
| | Neuroscientist |
| | Professor |
| | Other |
| | Are you a clinical scientist (both working as a clinician and doing research)? |
| **Inclusion criteria** | When considering the last ten years, how many years of experience do you have with the assessment and/or treatment of patients with disorders of consciousness (DoC)? |
| | When considering the last ten years, how many years of experience do you have in research on patients with DoC? |
| | How many articles (available on Pubmed) did you publish on DoC in the last 5 years? |
| **Setting** | Acute |
| | Rehabilitation (inpatient and/or outpatient) |
| | Long-term care (nursing home, skilled nurse facility - SNF, long term acute care - LTAC) |
| | Other |
| **Population** | Pediatric (< 18yo) |
| | Adult (18-64yo) |
| | Elderly (65yo and above) |
| **Technique access** | Behavioral |
| | Electroencephalogram–EEG |
| | Event-related potentials–ERP |
| | Functional magnetic resonance imaging – fMRI |

| | |
|---|---|
| | Positron emission tomography–PET |
| | Single-photon emission computed tomography–SPECT |
| | Transcranial magnetic stimulation combined with EEG–TMS-EEG |
| | Brain computer interface–BCI |
| | N/A |
| | Other |
| **Technique paradigm** | Passive listening |
| | Active tasks |
| | Rest |
| | N/A |
| | Other |
| **Before answering the survey, to what extent do you agree with the following name(s)** | |
| Covert awareness (CA) | |
| Functional locked-in (fLIS) | |
| Non-behavioral MCS (MCS*) | |
| Cognitive motor dissociation (CMD) | |
| *Comments:* | |
| **To what extent do you agree with parts of the definition below?** | |
| ***CA definition*** | non-communicative patients |
| | including those diagnosed as vegetative, minimally conscious, or locked in |
| | who may be able to use their residual cognitive capabilities |
| | to communicate their thoughts to those around them |
| | by modulating their own neural activity |
| | *General comments on the name Covert Awareness (CA)* |
| ***fLIS definition*** | a dissociation between motor dysfunctions and preserved higher cognitive functions |
| | as shown by functional imaging techniques |
| | and a consistent and reliable communication |
| | using non-speech and non-gestural communication comments |
| | through direct brain signaling |
| | *General comments on the name Functional Locked-In Syndrome (Functional LIS)* |

| | |
|---|---|
| ***MCS* definition*** | patients who are diagnosed as being in a vegetative state (VS; or unresponsive wakefulness syndrome - UWS) by bedside testing |
| | but then diagnosed MCS with neuroimaging techniques comments |
| | using passive or active paradigms comments |
| | *General comments on the name Non-behavioral Minimally Conscious State (MCS*)* |
| ***CMD definition*** | severely brain-injured patients |
| | including patients in vegetative state (VS; or unresponsive wakefulness syndrome - UWS), in minimally conscious state minus (or MCS-; i.e., without reproducible response to command), or complete Locked In Syndrome (LIS) |
| | who show a dissociation of a retained but unrecognized (covert) cognitive capacity |
| | *General comments on the name Cognitive Motor Dissociation (CMD)* |
| **After answering the survey, to what extent do you agree with the following name(s)** | |
| Covert awareness (CA) | |
| Functional locked-in (fLIS) | |
| Non-behavioral MCS (MCS*) | |
| Cognitive motor dissociation (CMD) | |
| *Comments* | |
| **Round 2** | |
| **Before answering the survey, to what extent do you agree with the following name(s)** | |
| Covert awareness (CA) | |
| Functional locked-in (fLIS) | |
| Non-behavioral MCS (MCS*) | |
| Cognitive motor dissociation (CMD) | |
| *Comments* | |
| **To what extent do you agree with parts of the new definition below?** | |
| ***CA definition*** | patients who are in a disorder of consciousness and who do not show functional communication verbally or by gestures |
| | including those behaviorally diagnosed as in a Vegetative State/Unresponsive Wakefulness syndrome |
| | to follow command and/or communicate |
| | *General comments on the name Covert Awareness (CA)* |

| | |
|---|---|
| ***fLIS definition*** | as shown by functional neuroimaging and/or electrophysiology |
| | and a reliable communication |
| | through brain modulation |
| | *General comments on the name Functional Locked-In Syndrome (Functional LIS)* |
| ***MCS* definition*** | patients who are initally diagnosed as being in a vegetative state (VS; or unresponsive wakefulness syndrome - UWS) by behavioral assessment |
| | but whose brain activity is similar to Minimally Conscious State (MCS) with neuroimaging and/or electrophysiology |
| | using active tasks |
| | *General comments on the name Non-behavioral Minimally Conscious State (MCS*)* |
| ***CMD definition*** | including patients behaviorally diagnosed as vegetative state (VS; or unresponsive wakefulness syndrome - UWS) or minimally conscious state minus (or MCS-; i.e., without reproducible response to command) |
| | who show a dissociation of a retained but unrecognized (covert) consciousness using active neuroimaging and/or electrophysiological paradigms |
| | *General comments on the name Cognitive Motor Dissociation (CMD)* |
| **After answering the survey, to what extent do you agree with the following name(s)** | |
| Covert awareness (CA) | |
| Functional locked-in (fLIS) | |
| Non-behavioral MCS (MCS*) | |
| Cognitive motor dissociation (CMD) | |
| *Comments* | |
| **To what extent do you agree with the following "covert" related name(s)?** | |
| Covert awareness | |
| Covert consciousness | |
| Covert cognition | |
| *Comments* | |

**eTable 2.** Definition statements with agreement rates for CA, CMD, MCS*, fLIS, according to geographic location (EU/UK, USA, Others) and background (neuroscientist)

| **EU/UK** | ***Mdn*** | ***IQR*** | ***n*** | ***% agree*** | ***Consensus*** |
|---|---|---|---|---|---|
| patients who are in a disorder of consciousness and who do not show functional communication verbally or by gestures | 5 | 1 | 36 | 89% | **y** |
| including those behaviorally diagnosed as in a Vegetative State/Unresponsive Wakefulness syndrome | 5 | 1 | 36 | 81% | **y** |
| to follow command and/or communicate | 5 | 0 | 36 | 81% | **y** |
| **USA** | | | | | |
| patients who are in a disorder of consciousness and who do not show functional communication verbally or by gestures | 5 | 2 | 23 | 74% | n |
| including those behaviorally diagnosed as in a Vegetative State/Unresponsive Wakefulness syndrome | 5 | 1 | 23 | 78% | **y** |
| to follow command and/or communicate | 5 | 2 | 23 | 74% | n |
| **Others** | | | | | |
| patients who are in a disorder of consciousness and who do not show functional communication verbally or by gestures | 5 | 1 | 15 | 87% | **y** |
| including those behaviorally diagnosed as in a Vegetative State/Unresponsive Wakefulness syndrome | 5 | 1 | 15 | 80% | **y** |
| to follow command and/or communicate | 5 | 1 | 15 | 87% | **y** |
| **Neuroscientist** | | | | | |
| patients who are in a disorder of consciousness and who do not show functional communication verbally or by gestures | 5 | 1 | 17 | 94% | **y** |
| including those behaviorally diagnosed as in a Vegetative State/Unresponsive Wakefulness syndrome | 5 | 0 | 17 | 76% | **y** |
| to follow command and/or communicate | 5 | 0 | 17 | 88% | **y** |

***Legend***: Covert Awareness (CA), Cognitive Motor Dissociation (CMD), Non-behavioral MCS (MCS*), functional Locked-In Syndrome (fLIS), Median (Mdn), Interquartile range (IQR), sample size (n), Consensus (yes/y, no/n)

**eTable 3.** Agreement rates for names such as CA, CMD, MCS*, fLIS as well as for covert related names, according to geographic location (EU/UK, USA, Others) and background (neuroscientist)

| EU/UK | *Mdn* | *IQR* | *n* | *% agree* | *Consensus* |
|---|---|---|---|---|---|
| **After taking the survey (Round 2)** | | | | | |
| Covert Awareness (CA) | 5 | 1 | 34 | 94% | y |
| Cognitive Motor Dissociation (CMD) | 4 | 2 | 34 | 74% | n |
| Non-behavioral MCS (MCS*) | 2 | 2 | 34 | 35% | n |
| functional Locked-In Syndrome (fLIS) | 2 | 1 | 34 | 6% | y |
| **Covert related names** | | | | | |
| Covert awareness | 4 | 1 | 34 | 88% | n |
| Covert consciousness | 4 | 3 | 34 | 65% | n |
| Covert cognition | 2 | 2 | 34 | 29% | n |
| **USA** | | | | | |
| **After taking the survey (Round 2)** | | | | | |
| Covert Awareness (CA) | 5 | 1 | 22 | 82% | y |
| Cognitive Motor Dissociation (CMD) | 4 | 2 | 22 | 73% | n |
| Non-behavioral MCS (MCS*) | 2 | 2 | 22 | 27% | n |
| functional Locked-In Syndrome (fLIS) | 2 | 2 | 20 | 20% | n |
| **Covert related names** | | | | | |
| Covert awareness | 4 | 1 | 22 | 77% | n |
| Covert consciousness | 5 | 2 | 22 | 73% | n |
| Covert cognition | 3 | 2 | 22 | 45% | n |
| **Others** | | | | | |
| **After taking the survey (Round 2)** | | | | | |
| Covert Awareness (CA) | 5 | 0 | 15 | 80% | y |
| Cognitive Motor Dissociation (CMD) | 4 | 1 | 15 | 73% | n |
| Non-behavioral MCS (MCS*) | 2 | 2 | 15 | 20% | n |
| functional Locked-In Syndrome (fLIS) | 2 | 2 | 15 | 13% | n |

| | | | | | |
|---|---|---|---|---|---|
| **Covert related names** | | | | | |
| Covert awareness | 5 | 1 | 15 | 80% | y |
| Covert consciousness | 4 | 2 | 15 | 60% | n |
| Covert cognition | 3 | 2 | 15 | 27% | n |
| **Neuroscientist** | | | | | |
| **After taking the survey (Round 2)** | | | | | |
| Covert Awareness (CA) | 5 | 1 | 17 | 82% | y |
| Cognitive Motor Dissociation (CMD) | 4 | 1 | 17 | 76% | n |
| Non-behavioral MCS (MCS*) | 2 | 3 | 17 | 47% | n |
| functional Locked-In Syndrome (fLIS) | 2 | 1 | 17 | 6% | y |
| **Covert related names** | | | | | |
| Covert awareness | 5 | 1 | 17 | 88% | y |
| Covert consciousness | 4 | 2 | 17 | 53% | n |
| Covert cognition | 2 | 2 | 17 | 29% | n |

***Legend***: Covert Awareness (CA), Cognitive Motor Dissociation (CMD), Non-behavioral MCS (MCS*), functional Locked-In Syndrome (fLIS), Median (Mdn), Interquartile range (IQR), sample size (n), Consensus (yes/y, no/n)
